# Well-being outcomes of a family-focused intervention for persons with type 2 diabetes and support persons: Main, mediated, and subgroup effects from the FAMS 2.0 RCT

**DOI:** 10.1101/2023.09.11.23295375

**Authors:** McKenzie K. Roddy, Andrew J. Spieker, Lyndsay A Nelson, Robert A. Greevy, Lauren M. LeStourgeon, Erin M. Bergner, Merna El-Rifai, Tom A. Elasy, James E. Aikens, Ruth Q. Wolever, Lindsay S. Mayberry

## Abstract

**Aims:** Type 2 diabetes self-management occurs within social contexts. We sought to test the effects of Family/friends Activation to Motivate Self-care (FAMS), a self-care support intervention delivered via mobile phones, on psychosocial outcomes for persons with diabetes (PWDs) and their support persons.

**Methods:** PWDs had the option to enroll with a friend/family member as a support person in a 15-month RCT to evaluate FAMS versus enhanced usual care. FAMS included 9-months of monthly phone coaching and text message support for PWDs, and text message support for enrolled support persons.

**Results:** PWDs (N=329) were 52% male and 39% from minoritized racial or ethnic groups; 50% enrolled with elevated diabetes distress. Support persons (N=294) were 26% male and 33% minoritized racial or ethnic groups. FAMS improved PWDs’ diabetes distress (*d*=-0.19) and global well-being (*d*=0.21) during the intervention, with patterns of larger effects among minoritized groups. Post-intervention and sustained (15-month) improvements were driven by changes in PWDs’ self-efficacy, self-care behaviors, and autonomy support. Among support persons, FAMS improved helpful involvement without increasing burden or harmful involvement.

**Conclusions:** FAMS improved PWDs’ psychosocial well-being, with post-intervention and sustained improvements driven by improved self-efficacy, self-care, and autonomy support. Support persons increased helpful involvement without adverse effects.

## 1. Introduction

The impact of type 2 diabetes is difficult to overstate, with estimated US prevalence of 9.3% in 2020 [1] and increasing rapidly [2]. Beyond physical consequences, a recent meta-analysis estimated 36% of persons with diabetes (PWDs) experience significant emotional distress about diabetes [3]. This diabetes distress is distinct from depression or generalized life stress [4,5] and associated with worse treatment adherence and worse glycemic management [6,7]. Notably, diabetes distress disproportionately impacts PWDs who are younger, female, depressed, and from minoritized racial and ethnic groups [3,8,9]. Furthermore, the trajectories of diabetes distress are more severe and persistent for PWDs with the lowest social support [10].

Diabetes self-management largely occurs in a social context [11–13]. Observational data has highlighted the importance of involvement from family and close friends. PWDs who report harmful family/friend involvement tend to show greater diabetes distress [14,15]. In turn, about 40% of cohabitating family members also experience clinically significant diabetes distress about their loved one’s diabetes [16]. Family/friends who have higher diabetes distress also exhibit more harmful involvement in the PWDs’ self-management [14]. Moreover, increased diabetes distress among family/friends is associated with increased diabetes distress among the PWDs [14]. There is a notable gap in research on diabetes distress and engaging family/friends; Sturt et al.’s review on interventions to reduce diabetes distress found only one study that included family members of adults with type 1 diabetes [17], and no studies of adults with type 2 diabetes. In a review of family interventions for adults with diabetes, Baig et al. found only four of 26 studies measured any outcomes for family members, and none assessed effects on family members’ distress [18]. Despite recommendations from the American Diabetes Association to incorporate family [19], very little is known regarding interventions that include family when targeting diabetes distress.

Individually delivered psychosocial and psychoeducational interventions for PWDs have demonstrated some success in reducing diabetes distress [17,20], with minimal differences between face-to-face and remote delivery [17]. More intensive interventions are associated with significant reductions in distress [20]; however, there is a paucity of data on diabetes distress interventions when family/friends are engaged. Few family interventions from adults with diabetes report on diabetes distress or quality of life [18]. Baig et al. identified four studies reporting these outcomes; of those, one found improvements in diabetes distress [21] and three reported improvements in quality of life [22–24]. All three studies reporting quality of life improvements were conducted within minoritized racial and ethnic groups, and they varied widely in intensity of family involvement in the intervention. Only one study reported measures of family/friend involvement and found trends towards increasing helpful (p=0.07) and harmful (p=0.07) involvement [23]. No study has examined whether changes in helpful or harmful family/friend involvement led to improvements in psychosocial outcomes for PWDs.

The current study presents results from a randomized controlled trial (RCT) evaluating a family-focused self-care support intervention, Family/friends Activation to Motivate Self-care (FAMS), on psychosocial aspects including diabetes distress and global well-being. We previously developed and piloted FAMS [25,26], and findings from the pilot were used to inform intervention improvements prior to evaluation in this FAMS 2.0 RCT [27]. FAMS is a mobile phone-delivered intervention (including phone calls and text message support) with the goal of improving PWDs’ self-efficacy, self-care behaviors, and family/friend involvement without increasing support person burden. Enrolled PWDs were given the option to co-enroll a support person (friend/family member) who also received text messages if the PWD was randomized to intervention (25). All analyses and outcomes were planned *a priori* [27]. Herein, we had three complementary aims. First, we examined the effects of FAMS on PWDs’ diabetes distress and global well-being during the 9-month intervention and sustained (15-month) effects. We also explored if there were differential patterns of effects by PWDs’ gender, race and ethnicity, socioeconomic status, and whether they were cohabitating with their support person. Second, we tested the hypothesis that improvements in PWD’s diabetes distress and well-being were driven (mediated) by improvements in intervention targets: diabetes self-efficacy, self-care behaviors, and family/friend involvement. Third, among enrolled support persons we tested whether FAMS reduced diabetes distress and increased helpful involvement in the PWDs’ self-management, while not worsening harmful involvement nor support burden. We also explored possible differential patterns of effects for support persons who were non-male, by race and ethnicity, and who were not cohabitating with the PWD.

## 2. Subjects, Materials, and Methods

### 2.1. Participants

We recruited adults receiving outpatient care at Vanderbilt University Medical Center primary care clinics in middle Tennessee. Eligible PWDs were between 18 and 75 years of age, could speak and read English, community dwelling, diagnosed with type 2 diabetes, prescribed at least one daily diabetes medication, and owned a mobile phone. We excluded PWDs with indications of concurrent hospice or dialysis services, congestive heart failure, concurrent cancer treatment, pregnancy, dementia, or schizophrenia; or those who self-disclosed recent or ongoing emotional, physical, or sexual abuse. Eligible support persons needed to be at least 18 years old, speak and read English, and have a mobile phone separate from the PWD.

### 2.2. Procedures

Study procedures and intervention details were previously reported by Mayberry et al. 2022 [27]. Study procedures were approved by the Vanderbilt University Institutional Review Board (#190905) and this trial is registered (ClinicalTrials.gov-NCT04347291). Briefly, potentially eligible PWDs were identified through the electronic medical record and mailed recruitment letters requesting they express their interest in the study or opt out. Research staff enrolled participants by phone and asked eligible PWDs to identify a friend or family member to enroll with them as a support person, though not required. Research staff obtained verbal informed consent via phone after answering all questions about the study.

Enrolled PWDs were randomized 1:1 to intervention or enhanced care as usual (control) using an adaptively stratified randomization process to ensure balance on baseline outcomes of interest. PWDs were informed of their randomization assignment via phone. All participants received informational print materials about diabetes throughout the study and completed assessments at baseline, 6-, 9-, and 15-months. For 9-months, PWDs assigned to the FAMS intervention received monthly coaching and daily text message support, and their support persons (if enrolled) received text message support [25]. Monthly coaching consisted of behavioral goal setting, skill building around family/friend involvement in diabetes management specific to the selected goal, and setting a verbal contract to use the learned skill with a specific person (who may or may not be the enrolled support person). Text messages to PWDs helped them monitor their own diabetes self-management efforts and provided encouragement to engage friends/family in their self-care efforts. Text messages to support persons encouraged dialogue about and involvement in the PWD’s diabetes self-management.

### 2.3. Measures

Demographic information was collected via self-report at baseline. PWDs and support persons completed additional measures via surveys administered at baseline, 6-, 9-, and 15-months post-baseline. Responses were collected via an online REDCap link [28,29], mailed copy, or by phone, per their preference. Psychometric properties of the measures are detailed in **Supplementary Table S1.**

#### 2.3.1. Demographics

Race and ethnicity were assessed by participant selecting all racial and ethnic groups that apply, then operationalized as non-Hispanic white only vs. non-Hispanic Black only vs. other minoritized race or ethnicity (i.e., Hispanic) for subgroup analyses. Socioeconomic disadvantage for PWDs was calculated based on self-reported education less than or equal to a high school degree or GED, uninsured or public health insurance only, and/or annual household income <$50,000 USD. Gender was self-reported as male, female, or other gender and operationalized as male vs. non-male for subgroup analyses. Finally, we operationalized dyads as cohabitating vs. not for subgroup analyses.

#### 2.3.2. Outcomes

Secondary outcomes for the RCT included PWDs’ diabetes distress and global well-being. Diabetes distress was assessed with the five-item Problem Areas in Diabetes (PAID; α=0.90) [30], which has high internal reliability, concurrent validity [31] and discriminant validity [30]. Global well-being was assessed with the five-item World Health Organization–Five Well-Being Index (α=0.90) [32] which has high construct and predictive validity and is responsive and sensitive to change [33].

For support persons, outcomes included their own diabetes distress, support burden, and specific aspects of their involvement including helpful, harmful, and alignment between their current and desired involvement in the PWD’s diabetes care and diabetes feelings. We used several measures developed and benchmarked for family members by the Diabetes Attitudes, Needs, and Wishes Second Study (DAWN2) [34]. Support persons’ diabetes distress was assessed by the Family Members Problem Areas in Diabetes (PAID-FM; α=0.87) [34], and support burden was assessed by a single item from DAWN2 [34] (“How much of a burden is it for you to help manage [PWD’s] diabetes?”). Measures of support person’s involvement in the PWD’s self-management included report on their own involvement via the Family/friend Involvement in Adults’ Diabetes – family member version (FIAD-FM) [35] helpful and harmful scales (α_helpful_=0.87, α_harmful_=0.56), and alignment with diabetes care and with diabetes feelings. The 2 items from DAWN2 used to evaluate alignment asked, “Considering the future, how involved would you like to be in… [PWD’s] diabetes care?” and “… helping [PWD] deal with their feelings about diabetes?” [34]. Response options ranged from “much less involved” to “much more involved,” so we analyzed these items to capture movement toward the middle response “as involved as you are now” and report percent with alignment between desired and actual involvement.

#### 2.3.3. Mediators

For PWDs, mediators included measures of diabetes self-efficacy, self-care behaviors, and family/friend involvement. Diabetes self-efficacy was assessed by the Perceived Diabetes Self-Management Scale (α=0.87) [36]. Self-care behaviors included dietary behavior, physical activity, and medication adherence. Dietary behavior was assessed with two scales of the Personal Diabetes Questionnaire: Use of Information for Dietary Decision Making (α=0.81) and Problem Eating Behavior (α=0.81) [37]. Physical activity was assessed with average weekly MET (metabolic equivalent of task) minutes, assessed by an adapted version of the Rapid Assessment of Physical Activity [38], which queries days and minutes spent engaging in light, moderate, and vigorous activity. Medication adherence assessed by the Adherence to Refills and Medications Scale for Diabetes (α=0.82) [39], reverse coded such that higher scores indicate more adherence.

Family/friend involvement was assessed by three measures querying support received from numerous important others, including the support person (if enrolled). The Important Others Climate Questionnaire (α=0.90) assessed autonomy support [40,41]. The 4-item adapted Family Emotional Involvement and Criticism Scale (α=0.85) assessed perceived criticism [42,43]. Helpful and harmful aspects of received support were assessed by the Family/friend Involvement in Adults’ Diabetes (FIAD) [35]. Prior evidence indicates both helpful and harmful involvement must be included when estimating effects on outcomes due to potential suppression effects [35,44]. Therefore, we calculated a family involvement difference score for mediation models which represented the overall valence of involvement at each assessment (helpful – harmful involvement) with a positive difference score indicating more helpful than harmful involvement.

### 2.4. Analytic Plan

#### 2.4.1. Missingness

To address missing data, we used multiple imputation via chained equations with a total of M=500 iterations. One imputation model included baseline, 6-and 9-month data during the intervention and a second imputation model included baseline and 15-month data for sustained effects.

#### 2.4.2. Intervention effects for PWDs

We used generalized estimating equations (GEE) with a working-independence correlation structure and identity link to estimate effects of FAMS at 6-and 9-months. We adjusted for baseline insulin and adjusted for the baseline value of each respective outcome using a restricted cubic spline with three knots (chosen at the first, second, and third quartiles). We allowed a two-way interaction between time and FAMS, along with a two-way spline interaction between time and the baseline outcome. We obtained point estimates, 95% confidence intervals, and p-values for each of the 6-and 9-month effects, along with omnibus tests for joint 6-and 9-month effects based on a Wald test with two degrees of freedom. To evaluate sustained (15-month) effects of the FAMS intervention, we used a linear regression model analogous to that described for our analysis of 6-and 9-month outcomes, specifically including condition as the predictor of interest and adjusting for baseline insulin use and the baseline measurement of each respective outcome (via restricted cubic splines, as previously described).

#### 2.4.3. Subgroup effects for PWDs

We performed several subgroup analyses for PWDs, stratifying each outcome model by gender (male/non-male), race or ethnicity (non-Hispanic White, non-Hispanic Black, and a group that included both non-Hispanic other race and Hispanic), socioeconomic disadvantage (no/yes), and cohabitation status (among those with enrolled support person: non-cohabitating/cohabitating).

#### 2.4.4. Mediation Analyses for PWDs

Mediation models were estimated using path analysis in Amos version 29 using the regression imputation approach to address missing data. We then used imputed data and 2,000 bootstrap samples to estimate bias-corrected 95% confidence intervals for estimates and indirect effects. For each outcome, we ran mediation models including condition as predictor, all hypothesized mediators at both 6-and 9-months, and the outcome of interest, adjusted for baseline values of the outcome of interest, and baseline values of each included mediator. Adjustment for baseline values aids with estimate precision and interpretability of effects in terms of changes in the outcomes of interest. Error variances were permitted to covary for the 6-and 9-month assessments of each mediator, and between all mediators as assessed at the same timepoint.

First, we ran models with all mediators. Then, to develop more parsimonious models, we included any mediator with *p*≤0.20 associated with the specific indirect effect at 6-or 9-months. Substantively, results between the full models and the more parsimonious models were consistent, but precision increased in the parsimonious models, so those are presented. Specific indirect effects presented represent the indirect effect via the mediator at both 6-and 9-months (i.e., summing the indirect effects via each time point).

#### 2.4.5. Intervention effects for Support Persons

To evaluate intervention effects on support person outcomes, we fit GEE models analogous to those described for PWD; because burden was measured on a 5-level scale, we did not feature a natural cubic spline but performed baseline adjustment with a single linear term. To evaluate sustained effects on the FAMS intervention (specifically, at 15 months), we used linear regression models analogous to those described for evaluating sustained effects for PWDs.

#### 2.4.6. Subgroup effects for Support Persons

We performed several subgroup analyses for support persons, stratifying our models for each outcome by gender (male/non-male), race or ethnicity (non-Hispanic White, non-Hispanic Black, and a group that included both non-Hispanic other race and Hispanic), and cohabitation status (non-cohabitating/cohabitating with PWD).

#### 2.4.7. Power

The study was designed with 80% power to detect standardized effect sizes of 0.32-0.34 with a retained sample size of 227-255 for intervention effects for PWDs and support persons [27]. All analyses used imputation for missing data and followed intention-to-treat principles, retaining all randomized participants and their support persons and therefore resulting in an effective sample size larger than that used in power analyses for both outcomes and mediation analyses.

## 3. Results

A total of 329 PWDs (294 with a co-enrolling support person) were randomized and participated longitudinally (total N=623). Demographic and baseline clinical characteristics for PWDs are in **Table 1** and support persons are in **Table 2**. All baseline standardized mean differences (SMDs) between FAMS and control were <0.20 for PWD except for lower income (SMD=0.35) and greater socioeconomic disadvantage (SMD=0.22) in FAMS. For support persons, all SMDs were <0.20 except lower income (SMD=0.31) and younger age (SMD=0.21) in FAMS. We retained at least 252 PWDs and 241 support persons at each timepoint (see **Supplementary Table S2**).

**Table 1:** Characteristics of Persons with Diabetes at Baseline.

|  | Mean±SD or n(%) |  |
| --- | --- | --- |
|  | Control (n=165) | FAMS (n=164) |
| *Demographic Characteristics* |  |  |
| Age, years | 57.9±10.5 | 56.1±11.0 |
| Gender, male | 85 (52%) | 84 (51%) |
| Race or ethnicity |  |  |
| Non-Hispanic white | 98 (59%) | 105 (64%) |
| Non-Hispanic Black | 42 (25%) | 35 (21%) |
| Non-Hispanic other race(s) | 12 (7%) | 10 (6%) |
| Hispanic | 11 (7%) | 13 (8%) |
| Socioeconomic status |  |  |
| Education, years | 15.3±3.0 | 15.4±2.8 |
| Annual household income, USD |  |  |
| <\$35,000 | 25 (15%) | 32 (20%) |
| \$35,000 - \$49,999 | 20 (12%) | 33 (20%) |
| \$50,000 - \$74,999 | 38 (23%) | 22 (13%) |
| \$75,000 - \$99,999 | 19 (6%) | 21 (13%) |
| ≥\$100,000 | 57 (35%) | 48 (29%) |
| Health insurance |  |  |
| Uninsured | 3 (2%) | 3 (2%) |
| Public only | 30 (18%) | 29 (18%) |
| Private | 129 (78%) | 126 (77%) |
| Socioeconomically disadvantaged | 67 (41%) | 85 (52%) |
| *Clinical Characteristics* |  |  |
| Diabetes duration, years | 11.7±7.7 | 11.3±8.5 |
| Medication regimen |  |  |
| Oral medications only | 107 (65%) | 97 (59%) |
| Oral medications and insulin | 47 (28%) | 55 (34%) |
| Insulin only | 9 (5%) | 9 (5%) |
| HbA1c % [mmol/mol] | 8.5±1.6 [69±17.5] | 8.6±1.8 [70±19.7] |
| *Intervention Targets* |  |  |
| Diabetes self-efficacy | 25.0±6.4 | 25.7±6.7 |
| Diabetes self-care |  |  |
| Problem eating behaviors | 3.8±1.0 | 3.7±1.1 |
| Use of dietary information | 3.1±1.5 | 3.0±1.5 |
| Physical activity | 830.1±968.7 | 670.4±840.3 |
| Medication adherence | 14.9±3.4 | 15.3±4.3 |
| Diabetes-specific family/friend involvement |  |  |
| Autonomy support | 3.5±0.9 | 3.4±0.8 |
| Perceived criticism | 3.5±3.4 | 4.2±4.1 |
| Involvement difference score | 0.60±1.01 | 0.67±1.01 |
| *Outcomes* |  |  |
| Psychosocial well-being |  |  |
| Diabetes distress | 38.1±24.5 | 38.8±26.9 |
| Global well-being | 56.5±23.1 | 55.6±23.4 |

*Note:* Family/friend Activation to Motivate Self-care (FAMS); United States Dollars (USD); Hemoglobin A1c (HbA1c).

**Table 2:** Characteristics of Support Persons at Baseline.

|  | Mean±SD or n(%) |  |
| --- | --- | --- |
|  | Control (n=144) | FAMS (n=150) |
| *Demographic Characteristics* |  |  |
| Age, years | 53.4±14.0 | 50.4±14.7 |
| Gender, male | 38 (26%) | 42 (28%) |
| Race or ethnicity |  |  |
| Non-Hispanic White | 88 (61%) | 93 (62%) |
| Non-Hispanic Black | 33 (23%) | 29 (19%) |
| Non-Hispanic other race(s) | 9 (6%) | 12 (8%) |
| Hispanic | 6 (4%) | 8 (5%) |
| Socioeconomic status |  |  |
| Education, years | 14.9±2.6 | 14.9±2.4 |
| Annual household income, USD |  |  |
| <\$35,000 | 18 (13%) | 25 (17%) |
| \$35,000 - \$49,999 | 15 (10%) | 21 (14%) |
| \$50,000 - \$74,999 | 28 (19%) | 22 (15%) |
| \$75,000 - \$99,999 | 20 (14%) | 24 (16%) |
| ≥\$100,000 | 52 (36%) | 41 (27%) |
| *Relationship Characteristics* |  |  |
| Relationship type |  |  |
| Spouse/partner of PWD | 85 (59%) | 85 (57%) |
| Parent of PWD | 15 (10%) | 15 (10%) |
| Son/daughter of PWD | 15 (10%) | 22 (15%) |
| Grandchild of PWD | 10 (6%) | 7 (5%) |
| Friend of PWD | 12 (7%) | 11 (7%) |
| Other | 5 (3%) | 4 (3%) |
| Cohabiting with PWD | 103 (72%) | 105 (70%) |
| *Outcomes* |  |  |
| Diabetes distress | 31.4±21.6 | 34.2±22.2 |
| Support burden | 0.4±0.7 | 0.4±0.8 |
| Diabetes-specific family/friend involvement |  |  |
| Helpful involvement | 2.5±1.0 | 2.7±0.9 |
| Harmful involvement | 1.5±0.5 | 1.5±0.4 |
| Involvement alignment in diabetes care [n aligned] | 2.9±0.8 | 2.9±0.9 |
| Involvement alignment with diabetes feelings [n aligned] | 2.9±0.9 | 3.0±0.9 |
| *Note:* Family/friend Activation to Motivate Self-care (FAMS); United States Dollars (USD); Person with diabetes (PWD). |  |  |

### 3.1. Outcomes for Persons with Diabetes

#### 3.1.1 PWD Intervention Effects

Among PWDs, there was a beneficial intervention effect on diabetes distress at 9-months (b=-4.83, 95% CI [-9.34, -0.32], *p*=0.036, Cohen’s *d*=-0.19), and a beneficial intervention effect for global well-being at 6-months (b=4.77, 95% CI [0.36, 9.18], *p*=-0.034, Cohen’s *d*=0.21; **Table 3**); however, the omnibus tests for diabetes distress (*p*=0.10) and global well-being (*p*=0.11) with all three timepoints did not reach significance during the intervention. We did not find evidence of sustained intervention effects at 15 months on diabetes distress or global well-being for PWDs (all *p*-values>0.40).

**Table 3:** Effects of FAMS on Outcomes for Persons with Diabetes (PWDs) and Support Persons.

| Outcome | 6 months |  | 9 months |  | Omnibus | 15 months |  |
| --- | --- | --- | --- | --- | --- | --- | --- |
|  | Est. (95% CI) | *p* | Est. (95% CI) | *p* | *p* | Est. (95% CI) | *p* |
| *PWDs Outcomes* |  |  |  |  |  |  |  |
| Diabetes distress | -3.35 (-7.99, 1.29) | 0.16 | -4.83 (-9.34, -0.32) | **0.036** | 0.10 | -2.01 (-6.95, 2.94) | 0.43 |
| Global well-being | 4.77 (0.36, 9.18) | **0.034** | 2.19 (-1.95, 6.33) | 0.30 | 0.11 | -1.78 (-6.32, 2.77) | 0.44 |
| *Support Person Outcomes* |  |  |  |  |  |  |  |
| Diabetes distress | 0.60 (-3.18, 4.38) | 0.76 | 0.03 (-3.77, 3.84) | 0.99 | 0.94 | -0.05 (-4.09, 3.97) | 0.98 |
| Support burden* | -0.04 (-0.20, 0.11) | 0.58 | 0.01 (-0.15, 0.16) | 0.91 | 0.80 | -0.07 (-0.21, 0.07) | 0.33 |
| Helpful family/friend involvement | 0.42 (0.24, 0.59) | **<0.001** | 0.41 (0.22, 0.58) | **<0.001** | **<0.001** | 0.83 (-0.70, 2.35) | 0.29 |
| Harmful family/friend involvement | -0.01 (-0.09, 0.07) | 0.76 | -0.01 (-0.09, 0.07) | 0.82 | 0.95 | 0.01 (-0.07, 0.09) | 0.84 |
| Alignment-care** | 0.04 (-0.12, 0.20) | 0.60 | 0.00 (-0.15, 0.15) | 0.99 | 0.86 | 0.06 (-0.11, 0.23) | 0.50 |
| Alignment-feelings** | 0.01 (-0.17, 0.18) | 0.95 | 0.02 (-0.15, 0.19) | 0.80 | 0.97 | 0.05 (-0.11, 0.22) | 0.53 |

*Note:* Bolded terms are significant at the $p < 0.05$ level. \*No spline included (only a five-category variable). \*\*Transformed as absolute value of (X-2); negative indicates moving closer to two (better). Estimates for 6- and 9-months were obtained from a single longitudinal model and estimates for 15-months (post-intervention) were obtained from separate models.

#### 3.1.2 PWD Subgroup Analyses

For PWDs representing historically at-risk groups (non-males; group that included both non-Hispanic other race and Hispanic; socioeconomically disadvantaged), FAMS had a pattern of larger estimated effect sizes on global well-being. Further, compared to cohabitating dyads, PWDs in non-cohabitating dyads showed a pattern of larger estimated effect sizes in diabetes distress and global well-being (**Table 4**).

**Table 4:** Primary Outcome Estimates and 95% Confidence Intervals for Subgroups of Persons with Diabetes during Intervention.

| Outcomes | Gender |  | Race or Ethnicity |  |  | Disadvantaged |  | Cohabiting |  |
| --- | --- | --- | --- | --- | --- | --- | --- | --- | --- |
|  | Male | Non-male | Non-Hispanic white | Non-Hispanic Black | Other race(s) or ethnicities | No | Yes | No | Yes |
| *Diabetes Distress* |  |  |  |  |  |  |  |  |  |
| 6 months | -3.75<br>(-9.78, 2.29) | -3.30<br>(-10.40, 3.79) | -2.40<br>(-7.82, 3.02) | -8.31<br>(-18.90, 2.33) | 1.18<br>(-13.10, 15.50) | -4.59<br>(-10.80, 1.61) | -1.83<br>(-8.72, 5.06) | -6.14<br>(-15.10, 2.77) | -2.75<br>(-8.36, 2.86) |
| 9 months | -4.30<br>(-10.30, 1.67) | -5.18<br>(-11.80, 1.49) | -4.15<br>(-9.46, 1.14) | -6.48<br>(-16.60, 3.67) | -4.71<br>(-18.80, 9.40) | -5.26<br>(-11.60, 1.07) | -4.04<br>(-10.30, 2.18) | -8.25<br>(-16.90, 0.37) | -3.81<br>(-9.29, 1.68) |
| *Global Well-being* |  |  |  |  |  |  |  |  |  |
| 6 months | 3.32<br>(-2.85, 9.49) | 6.15<br>(-0.13, 12.40) | 3.45<br>(-1.96, 8.85) | 5.41<br>(-4.67, 15.50) | 11.70<br>(0.15, 23.30) | 2.15<br>(-3.25, 7.55) | 8.29<br>(1.11, 15.50) | 8.80<br>(-0.65, 18.30) | 3.25<br>(-1.68, 8.18) |
| 9 months | -1.00<br>(-6.55, 4.54) | 5.49<br>(-0.61, 11.60) | 0.87<br>(-4.07, 5.81) | 2.16<br>(-7.14, 11.50) | 9.20<br>(-1.89, 20.30) | 1.35<br>(-4.29, 7.00) | 3.27<br>(-2.81, 9.36) | 7.33<br>(-1.20, 15.90) | 0.53<br>(-4.20, 5.25) |

### 3.1.3 PWD Mediation Analyses

In mediation analyses (**Table 5**), there was also a total effect of FAMS on diabetes distress at 9 months (b=-4.52, *p*=0.017), comprised of a significant indirect effect (b=-5.15, 95% CI [-8.50, -2.60], *p*=0.001) and non-significant direct effect, indicating the benefits of FAMS for distress were driven by improvements in intervention targets. The specific intervention targets contributing most to driving indirect effects on distress at 9 months were improved self-efficacy and family/friend involvement difference (helpful-harmful). At 15 months, there was still a significant total indirect effect of FAMS on diabetes distress (b=-4.50, 95% CI [-7.69, -1.96], *p*=0.001), driven primarily by improvements in self-efficacy and problem eating behavior made during the intervention period.

**Table 5:**
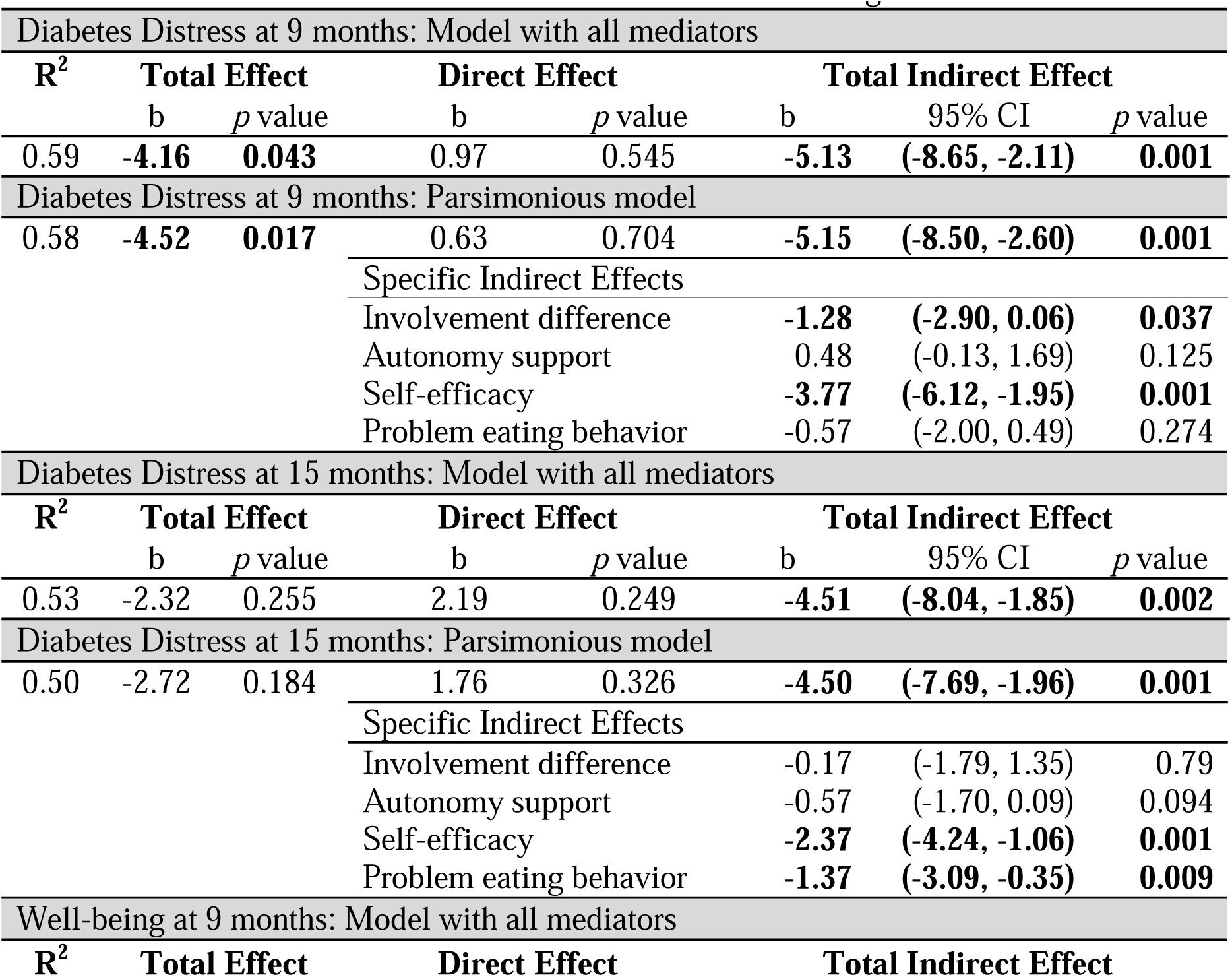
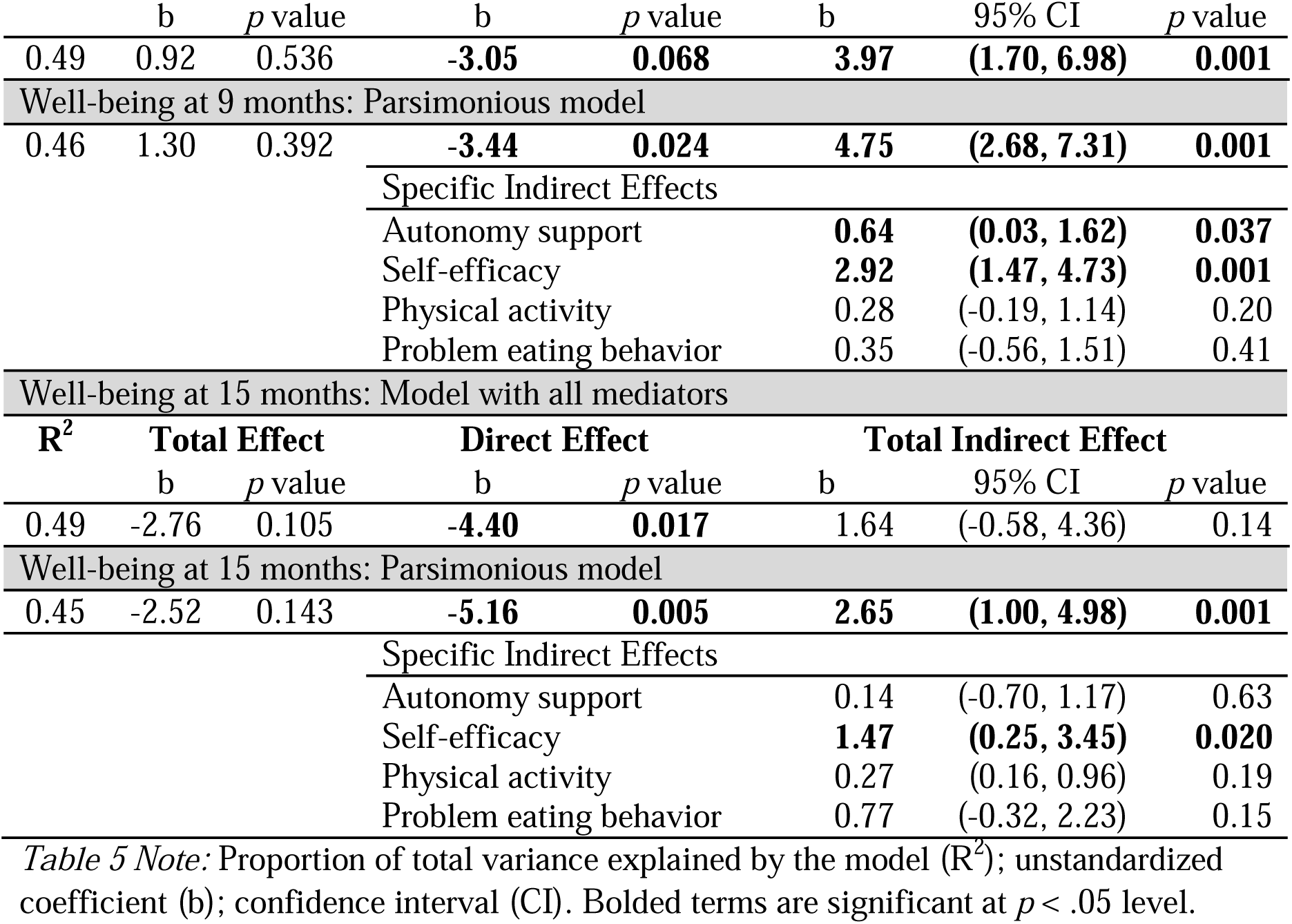
Mediation Models for Diabetes Distress and Well-being.

For global well-being, there was also a significant direct effect total indirect effect (b=2.68, 95% CI [2,68, 7.31], *p*=0.001) of FAMS on well-being at 9 months which was sustained at 15-months (b=2.65, 95% CI [1.00, 4.98], *p*=0.001). The specific intervention targets driving indirect effects on well-being were improvements in autonomy support and self-efficacy. However, for global well-being we also found significant direct effects in the opposite direction. This indicates FAMS-related improvements in well-being were sustained for people with improvements in intervention targets but in the absence of sufficient improvements in intervention targets, global well-being worsened by 15 months for FAMS participants. This results in null total (mean) effects of FAMS on well-being.

### 3.3 Outcomes for Support Persons

#### 3.3.1 Support Person Intervention Effects

FAMS increased support persons’ helpful involvement (omnibus *p*<0.001) but did not significantly affect support persons’ harmful family involvement. FAMS did not reduce support persons’ diabetes distress but also did not increase support burden (see **Table 2**). The percentage of support persons reporting alignment between current and desired involvement increased in both study arms by 9 months (diabetes care: baseline 32% both arms to 52% control vs. 50% FAMS; diabetes feelings: 28% vs. 29% to 60% vs. 66%). We did not detect evidence of sustained intervention effects for support persons (all *p*-values>0.25).

#### 3.3.2 Support Person Subgroup Analyses

Effects of FAMS on diabetes distress and support burden for support persons did not indicate differential effects across tested groups by gender or race or ethnicity. There may have been a slight increase in distress for non-cohabitating support persons at 6-months, but this was no longer evident by 9-months (see **Supplementary Table S3**).

## 4. Discussion

FAMS improved PWDs’ diabetes distress and global well-being, but improvements were not robust across time, with early improvements in well-being and growing improvements in distress. Mediation analyses indicated PWDs who had sufficient improvements in intervention targets (particularly, family/friend involvement, autonomy support, self-efficacy, and problem eating behavior) during the intervention period had sustained improvements in diabetes distress and global well-being 6-months after the intervention ended. Our results contribute to literature describing interventions that improve diabetes distress and well-being for PWDs, including remotely delivered interventions [17]. This is the second intervention, to our knowledge, that was family-focused and targeted diabetes distress for PWDs. With Veterans, Aikens et al. found significant reductions in diabetes distress of a similar magnitude [21] from an intervention using technology to engage out-of-home support persons. In addition, all previous data on well-being improvements from family-focused interventions were gathered in minoritized racial or ethnic samples [22–24]; our work expands this to show positive impact on wider samples. Importantly though, FAMS had patterns of larger effects for lowering distress and improving well-being for historically at-risk groups. This is vital given that diabetes distress is more prevalent among females, younger individuals, and minoritized racial and ethnic groups [3,8,9]. Mediation models also revealed a detrimental direct effect of FAMS on PWDs’ well-being. This indicates that once the effect of FAMS on well-being through the mediators was accounted for, FAMS decreased well-being. It is possible that increasing focus on self-care and family/friend involvement diminished overall well-being for participants who did not experience improvements in those areas.

This work also adds to literature on family-focused intervention outcomes for enrolled friends/family members, as previous work only measured a single family-focused outcome or none at all [18]. FAMS increased helpful involvement from support persons without increasing burden or harmful involvement. It is critically important to strike a careful balance: asking family/friends to be more involved in helpful ways and avoiding furthering burdening them or inadvertently increasing harmful involvement [14,35,44]. It is not surprising that we did not detect improvements in support persons’ diabetes distress, as the intervention focused on PWDs and included only light touch intervention components for support persons. Finally, this is the first study to our knowledge to investigate differential patterns of effects for support persons by gender, race or ethnicity, and dyads’ cohabitation status. Given the increasing prevalence of long-distance caregiving relationships [45], it is useful that FAMS engages both cohabitating and non-cohabitating support persons – without evidence of harm or increased burden. FAMS thus offers wider generalizability than most family interventions for adults with chronic disease.

### 4.1 Strengths and Limitations

This trial had a robust methodological and statistical plan, increasing confidence that findings are not due to poor design or insufficient power. This study used self-report measures with their inherent balance of validity and ease to collect against their vulnerability to bias. For support persons, more intensive interventions components may be required for distress reduction; however, here is a tradeoff between greater intervention burden and larger effects. We tested some mediators of FAMS on distress and well-being, and their selection was guided by theory and prior work, but there may be meaningful mediators we did not assess or include in our analyses. The generalizability of these findings reflects the sample: English-speaking adults with type 2 diabetes receiving care at a large academic medical center in the mid-south United States.

### 4.2 Clinical Implications

In an RCT of the FAMS intervention, we found improvement in intervention targets were associated with improvements in distress and well-being. However, it is unclear whether PWDs who did not benefit need higher intensity interventions or ones that focus on areas other than self-care, family/friend involvement, and self-efficacy. For instance, some PWDs may benefit from more connection with clinical care to co-manage medication alongside behavioral changes, or more direct content on diabetes distress and coping skills. Further, we found evidence of potentially larger benefits of FAMS among historically at-risk groups. More work is needed to better understand who needs what type of intervention. Finally, FAMS improved helpful involvement of support persons, but other interventions may be better suited for support persons with elevated distress related to their loved one’s diabetes. Further work is needed to identify interventions that reduce diabetes distress among family members. In sum, the FAMS 2.0 RCT was a well-designed and executed trial of a family-focused self-care support intervention for adults with T2D that found modest improvements in well-being for PWDs and improvements in helpful involvement without increasing harmful involvement or support burden for support persons.

## Data Availability

All data produced in the present study are available upon reasonable request to the authors

## Acknowledgements

The study was funded by the National Institute of Diabetes and Digestive and Kidney Diseases (R01DK119282). MK Roddy was supported by the Vanderbilt Faculty Research Scholars. An abstract accepted for the 44^th^ Annual Meeting and Scientific Sessions of the Society of Behavioral Medicine was published in the *Annals of Behavioral Medicine* 2023; 57 (Suppl. 1):S292.

## Declaration of competing interests

The authors declare that they have no known competing financial interests or personal relationships that could have appeared to influence the work reported in this paper.

## CRediT authorship contribution statement

**McKenzie K. Roddy**: Formal analysis, Investigation, Writing - Original Draft, Supervision. **Andrew J. Spieker**: Data Curation, Formal analysis, Visualization, Writing - Review & Editing. **Lyndsay A. Nelson**: Methodology, Writing - Review & Editing. **Robert A. Greevy**: Methodology, Writing - Review & Editing, Supervision. **Lauren M. LeStourgeon**: Data Curation, Visualization, Project administration, Investigation, Writing - Review & Editing. **Erin M. Bergner**: Writing - Review & Editing, Supervision, Project administration. **Merna El-Rifai**: Project administration, Investigation, Writing - Review & Editing. **Tom A. Elasy**: Conceptualization, Methodology, Resources, Writing - Review & Editing. **James E. Aikens**: Methodology, Writing - Review & Editing. **Ruth Q. Wolever**: Supervision, Methodology, Writing - Review & Editing. **Lindsay S. Mayberry**: Conceptualization, Methodology, Writing - Review & Editing, Formal analysis, Resources, Supervision, Project Administration, Funding Acquisition.

## Notes

### Competing Interest Statement

The authors have declared no competing interest.

### Clinical Trial

NCT04347291

### Clinical Protocols

https://classic.clinicaltrials.gov/ProvidedDocs/91/NCT04347291/Prot_001.pdf

https://doi.org/10.1016/j.cct.2022.106956

### Author Declarations

The Institutional Review Board of Vanderbilt University gave ethnical approval for this work.

