## Supplementary for "Well-being outcomes of a family-focused intervention for persons with type 2 diabetes and support persons: Main, mediated, and subgroup effects from the FAMS 2.0 RCT"

| **Supplementary Table S1**: Measure Properties | | | | |
| --- | --- | --- | --- | --- |
| Construct | Measure | Description | Validity^1^ | Reliability^2^ |
| *Persons with Diabetes (PWD)* | | | | |
| Diabetes Distress | Problem Areas in Diabetes (PAID) [1] | Five-item measure assessing emotional, psychological, and social distress related to diabetes management. Responses range from 0=“not a problem” to 4=“serious problem.” Sum scores were transformed to range from 0 to 100 where higher scores indicate more distress. | Correlates with distress and HbA1c; negatively corelates with self-care behaviors [2] | α = 0.90 |
| Global Well-Being | World Health Organization – Five Well-Being Index (WHO-5) [3] | Five-item measure assessing overall emotional wellbeing. Responses range from 0=“at no time” to 5=“all of the time.” Scores range from 0 to 100 where higher scores indicate better global mental wellbeing. | High construct and predictive validity and screening instrument for depression [4] | α = 0.90 |
| Diabetes Self-Efficacy | Perceived Diabetes Self-Management Scale (PDSMS) [5] | Eight items assessing confidence in managing diabetes. Responses range from 1=“strongly disagree” to 5=“strongly agree.” Items reverse scored as appropriate and summed to produce a total score ranging from 8 to 40 with higher scores indicating better self-efficacy. | Correlated with self-reported self-care activities and measures of HbA1c [5] | α = 0.87 |
| Dietary Behavior | Personal Diabetes Questionnaire (PDQ) dietary behavior scales [6] | Problem Eating Behavior: Three items assessing overeating, unplanned snacks, and poor food choices. Use of Information for Dietary Decision Making: Three items assessing frequency of using information on the number of calories, carbohydrates, and grams of fat in foods to make decisions about what to eat.  For both scales, responses range from 1=“never” to 6=“1 or more times per day;” items reverse-scored as appropriate and averaged to create separate subscale scores ranging from 1 to 6. Higher scores indicate more problem eating behaviors and more use of dietary information for decision making. | Correlated with other self-report diet measures [6] and with HbA1c [7] | Problem Eating Behavior: α = 0.81  Information for Dietary Decision Making: α = 0.81 |
| Physical Activity | Rapid Assessment of Physical Activity (RAPA) [8] | Six items assessing number of days engaged in light, moderate, and vigorous physical activity and amount of time spent in each during a typical week. To increase sensitivity of the measure, we adapted the RAPA to calculate average weekly MET minutes using standard MET minutes for light (3.3), moderate (4) and vigorous activities (8). The equation summed the following for each category of activity: (number of days engaged) x (number of minutes engaged per time) x (MET minutes associated). Total scores range from 0 to 6533 with higher scores indicating more physical activity. | Correlated with caloric expenditure [8] | n/a |
| Medication Adherence | Adherence to Refills and Medications Scale for Diabetes (ARMS-D) [9] | Eleven items assessing adherence to diabetes medications. Responses range from 1= “none of the time” to 4=“all of the time”. Reverse-scored items as appropriate and summed to produce a total score ranging 11 to 44; we reversed the scale so higher scores indicate better adherence. | Validated against other self-report measures and objective refill adherence measures [10]; Independently predicts HbA1c [9] | α = 0.82 |
| Family/friend Involvement | Family/friend Involvement in Adults’ Diabetes (FIAD) [11] | Helpful scale: Nine items assessing helpful behaviors over the past month.  Harmful scale: Seven items assessing harmful behaviors over the past month.  Responses range from 1=“never in the past month” to 5=“twice or more each week”. Items are averaged with scales for scores ranging from 1 to 5. Higher helpful scale scores indicate more helpful behaviors from family/friends, and higher harmful scale scores indicate more harmful behaviors from family/friends. | Scales associated with self-care behaviors and HbA1c [11] | Helpful subscale: α = 0.89  Harmful subscale: α = 0.60 |
| Family/friend Involvement – Autonomy Supportive Communication | Important Others Climate Questionnaire [12,13] | Six items assessing family involvement specific to diabetes management. Responses range 1=“strongly disagree” to 5=“strongly agree.” Items averaged to create a total score ranging 1 to 5 with higher scores indicating more autonomy supportive communication. | Single factor distinguishable from motivation [12]; convergent validity with other measures of family/friend involvement [11] | α = 0.90 |
| Family/friend Involvement – Perceived Criticism | Adapted Family Emotional Involvement and Criticism Scale [14] | Four items assessing perceived specific to diabetes management. Responses range from 0=“almost never” to 4=“almost always”. Items summed to create a total score ranging from 0 to 16 with higher scores indicating more perceived criticism related to diabetes management. | Correlated negatively with communication, problem solving, cohesion, and adaptability [14]; convergent validity with other measures of family/friend involvement [15] | α = 0.85 |
| *Support Persons* | | | | |
| Diabetes Distress | DAWN2 Family Members Problem Areas in Diabetes (PAID-FM) [16] | Five-item measure assessing emotional, psychological, and social distress related to their loved one having diabetes. Responses range from 0=“not a problem” to 4=“serious problem.” Sum scores were transformed to range 0-100 where higher scores indicate more distress about the PWDs’ diabetes experienced by the support person. |  | α = 0.87 |
| Support Burden | DAWN2 Family Burden Item [16] | Single item that asks how much of a burden it is to help the PWD manage their diabetes, with responses on a scale from 0=“no burden” to 4=“a very large burden.” Higher scores indicate greater burden. |  | n/a |
| Family Involvement | Family/friend Involvement in Adults’ Diabetes (FIAD-FM) [11] | Helpful scale: Nine items assessing the support person’s own performance of helpful behaviors over the past month.  Harmful scale: Seven items assessing the support person’s own performance of harmful behaviors over the past month.  Responses range from 1=“never in the past month” to 5=“twice or more each week”. Reverse-scored items as appropriate and averaged with scales for scores ranging 1 to 5. Higher helpful scale scores indicate more helpful behaviors provided to the PWD, and higher harmful scale scores indicate more harmful behaviors provided to the PWD. | Two-factor solution identified in exploratory factor analysis [17]; associated with own diabetes distress [17] | Helpful α = 0.87  Harmful α = 0.56 |
| Alignment | 2 items from DAWN2 Family Experience of Patient Involvement measure [16] | Two items assessing alignment between desired and actual involvement with Diabetes Care and Feelings about Diabetes. Responses range 0=“much less involved” to 4=“much more involved” where a score of 2 indicates alignment between the support person's current level of involvement and their desired level of involvement. |  | n/a |

*Supplementary Table 1 Note*: α = alpha; PWD = person with diabetes; DAWN2 = Second Diabetes Attitudes, Wishes, and Needs Study; HbA1c, hemoglobin A1c; MET, metabolic equivalent of task; n/a, not applicable (indicated for single item measures or scales that include items assessing different information for which alpha is not applicable). ^1^Based on prior studies. ^2^Internal consistency of baseline measure in this study.

| **Supplementary Table S2**: Survey Completion Rates | | | | |
| --- | --- | --- | --- | --- |
|  | **Control** | | **FAMS** | |
|  | PWD (N=165) | SP (N=144) | PWD (N=164) | SP (N=150) |
| Mid-Intervention  (6 months) | 134 (81.2%) | 126 (87.5%) | 148 (93.3%) | 129 (86.0%) |
| Post-Intervention  (9 months) | 134 (81.2%) | 125 (86.1%) | 135 (85.4%) | 130 (86.7%) |
| Sustained  (15 months) | 121 (73.3%) | 117 (81.2%) | 131 (87.0%) | 124 (82.7%) |

*Supplementary Table 2 Note:* PWD = person with diabetes; SP = support person

| **Supplementary Table S3:** Estimates and 95% Confidence Intervals for Subgroups of Primary Outcomes during Intervention for Support Persons | | | | | | | | | | | | | | | | |
| --- | --- | --- | --- | --- | --- | --- | --- | --- | --- | --- | --- | --- | --- | --- | --- | --- |
|  |  | | Gender | | | | | Race or Ethnicity | | | | | Cohabitating | | | |
| Outcomes | | | Male | Non-male | | Non-Hispanic white | | | Non-Hispanic Black | | Other race(s) or ethnicities | | | No | | Yes |
| Diabetes Distress | |  | | |  | |  | | |  | |  | | |  | |
| 6 months | | | -0.96  (-8.35, 6.44) | 1.22  (-0.32, 5.63) | | -1.01  (-4.97, 2.96) | | | 2.67  (-4.80, 10.1) | | 0.96  (-8.85, 10.8) | | | 5.44  (-1.85, 12.7) | | -1.13  (-5.73, 3.08) |
| 9 months | | | 2.38  (-4.51, 9.27) | -0.85  (-5.33, 3.64) | | 0.163  (-4.56, 4.89) | | | -3.32  (-11.6, 4.94) | | 4.08  (-6.52, 14.7) | | | -1.11  (-8.31, 6.09) | | 0.32  (-4.11, 4.76) |
| Support Burden | |  | | |  | |  | | |  | |  | | |  | |
| 6 months | | | 0.15  (-0.17, 0.47) | -0.12  (0.30, 0.05) | | -0.09  (-0.25, 0.06) | | | -0.02  (-0.24, 0.21) | | 0.04  (-0.28, 0.37) | | | 0.05  (-0.16, 0.25) | | -0.10  (-0.30, 0.10) |
| 9 months | | | 0.13  (-0.14, 0.41) | -0.03  (-0.22, 0.15) | | -0.03  (-0.24, 0.17) | | | 0.18  (-0.09, 0.46) | | -0.07  (-0.52, 0.38) | | | 0.05  (-0.18, 0.28) | | -0.02  (-0.21, 0.17) |

*Supplementary Table 3 Note*: The table presents estimates and (95% confidence intervals).
